# Predicting Current Juul use among Emerging Adults through Twitter Feeds

**DOI:** 10.1101/19010553

**Authors:** Tung Tran, Melinda Ickes, Jakob W. Hester, Ramakanth Kavuluru

**Author notes:** **Corresponding Author** Address: 230E MDS Building, 725 Rose St., Lexington, KY 40508.

## Abstract

**Introduction:** Can we predict whether someone uses Juul based on their social media activities? This is the central premise of the effort reported in this paper. Several recent social media-related studies on Juul use tend to focus on the characterization of Juul-related messages on social media. In this study, we assess the potential in using machine learning methods to automatically identify whether an individual uses Juul (past 30-day usage) based on their Twitter data.

**Methods:** We obtained a collection of 588 instances, for training and testing, of Juul use patterns (along with associated Twitter handles) via survey responses of college students. With this data, we built and tested supervised machine learning models based on linear and deep learning algorithms with textual, social network (friends and followers), and other hand-crafted features.

**Results:** The linear model with textual and follower network features performed best with a precision-recall trade-off such that precision (PPV) is 57% at 24% recall (sensitivity). Hence, at least every other college-attending Twitter user flagged by our model is expected to be a Juul user. Additionally, our results indicate that social network features tend to have a large impact (positive) on predictive performance.

**Conclusion:** There are enough predictive signals from social feeds for supervised modeling of Juul use, even with limited training data, implying that such models are highly beneficial to very focused intervention campaigns. Moreover, this initial success indicates potential for more involved automated surveillance of Juul use based on social media data, including Juul usage patterns, nicotine dependency, and risk awareness.

## 1. Introduction

Juul remains the most popular and well-known brand of e-cigarette, especially among teens and emerging adults, encompassing 72% of the market share as of August 2019. While sales campaigns claim they are not specifically targeted at minors, Juul use has become popular among teenagers, prompting public health concerns and subsequent ongoing investigations by the U.S. Food and Drug Administration [1,2]. The popularity of Juul use among young people is partly facilitated by its resemblance to a USB-drive, high level of nicotine, and emissions that are hard to see [3]. Several recent studies explore and characterize social media posts related to Juul use and dependency [4,5,6,7]; however, to our knowledge, none are designed for the purpose of predictive modeling of Juul usage by consumers based on their social media feeds. In this first of its kind study, using self-reported survey answers as ground truth, we explore the potential for using publicly available historical Twitter data, including Twitter messages (tweets) and friend and follower networks, to predict an individual’s past 30-day use of Juul. We compare supervised machine learning models based on traditional linear and more recent deep learning algorithms and identify influential features for this predictive task. We contend that models that are able to automatically distinguish between users and non-users of Juul in a quick and inexpensive manner will benefit highly focused interventional campaigns on social media. In the end, we imagine this potential application to be complementary to more field level conventional interventions and thus believe our proposed idea will be part of a multi-pronged approach in tobacco prevention and control.

## 2. Materials and Methods

### 2.1. Data Collection

The dataset is derived from a longitudinal study on Juul use among incoming college students (an important emerging adult subgroup) at the University of Kentucky conducted between August 16, 2018 and April 30, 2019. Participants were invited to participate in an online survey and asked various questions regarding patterns of Juul use, dependency, and general demographic information over three time points (T1, T2, and T3) spaced approximately three to five months apart. During T1, 555 participants optionally provided their Twitter handle (i.e., user name or identification). Of these, 214 participated again at T2 and 170 again participated at T3. Hence, we have 939 total discrete survey records that are each associated with a Twitter handle. As tweets are an essential predictive feature, we keep only records associated with at least one tweet in the 90-day window immediately prior to the day the survey response was received. The final dataset consisted of 588 records. We observed answers to the survey question on whether the participant has used Juul in the past thirty days as a discrete binary Yes/No answer in the ground truth. Of the 588 instances in the dataset, 151 (25.6%) reported current Juul use and were categorized as Juul users.

### 2.2. Evaluation Procedure

We contend that precision-focused metrics are reasonable for assessing model performance in terms of practical utility. This is especially true in the context of highly focused interventions where it is more important to ensure resources are prioritized for people who are highly likely to be Juul users. While coverage (sensitivity) is important, there is no interventional expense associated with false negative errors. Thus, we evaluated the proposed models with precision-focused metrics. One choice of metrics is *F*_β_ with 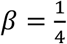, which is the harmonic mean between precision and recall with a greater emphasis on precision. Another metric used was *average precision at* ***n*** [8], a metric commonly used in information retrieval tasks for search engines. As commonly applied, we selected **n** = 10 and normalized by **n**. Intuitively, suppose we ranked all test examples by predicted likelihood of being a Juul user; average precision at 10 (*AP*@10) measures the quality of results in the top 10 considering the number of relevant results (i.e., positive for Juul use) and their placement in the ranking. We note that *F*_β_is more interpretable in terms of precision-recall trade-off, while *AP*@10 is useful for fine-grained comparisons. We performed our experiments using a *stratified* 5-fold cross-validation setup repeated **six** times. On each rotation, we reserved one fold for testing, one fold for validation, and the remaining (three) folds for training. Overall model performance is assessed based on the average of the aforementioned metrics over the 30 fold-wise evaluations.

### 2.3. Machine Learning Models

#### Traditional Linear Model

The well-known logistic regression algorithm with bag-of-word features served as our linear model. We concatenated all tweets and encode each *n*-gram (for *n* = 2, …, 6) as a 0/1 binary feature (ignoring frequency). We ignored *n*-grams with a global frequency less than three. These settings were chosen based on preliminary experiments. During training, we optimized the regularization hyperparameter *c* and threshold *t*, by performing grid search over the held-out validation fold, such that *F*_β_ is maximized. In a typical binary classification model, *t* = 0.5 and examples with *p*(*x*) > *t* are labeled positive; however, varying *t* allows us to obtain a desirable trade-off between precision and recall as determined by the *F*_β_ metric. Prior to testing, we trained on both the training and validation set using the optimized *c* value and predicted as positive only test examples where *p*(*x*) > *t*.

#### Deep Learning Model

Our deep learning model is based on an LSTM-based recurrent neural network [9] over the list of tweets in chronological order. Each tweet is first encoded as a 100-dimensional vector using a standard convolutional neural network [10] of window sizes 2, 3, and 4 with 50 filters each that convolves over randomly initialized word vectors of size 200. Our preliminary experiments showed that these settings were optimal, and inclusion of pretrained embeddings resulted in worse model performance. Once tweets are processed by the recurrent network, the output state at the last recurrent unit is fully connected to a softmax output layer for predicting Juul use.

### 2.4. Feature Extraction

#### Textual features

For both linear and deep models, we used the NLTK Twitter-aware tokenizer [11], designed for tokenizing tweets, that takes into consideration domain-specific tokens such as smileys (stylized faces constructed using punctuation symbols) and hashtags (user-generated metadata tag). Only tweets made within a 90-day window immediately prior to survey response were kept for prediction. Both linear and deep learning models effectively treated input text as n-gram based features.

#### User features

We included features specific to the Twitter account including the age of the account, number of tweets, whether the user is verified, number of followers, number of friends, and number of lists the user is assigned. For both linear and deep learning models, these features were encoded as vectors and are concatenated to the n-gram feature vector immediately prior to the softmax layer.

#### Tweet features

We additionally included features specific to a tweet which include the age of the tweet, whether the tweet is a reply, a retweet, or a quote, and the number of times the tweet has been retweeted. As the linear model does not encode tweets individually, but rather as a singular “blob” of text, tweet features were only included in the deep learning model.

#### Friend and follower network

Lastly, we included social network-based features based on friends and followers associated with the account. ‘Followers’ are a list of users that are subscribed to one’s tweets, while ‘friends’ are a list of users to which one is subscribed. Intuitively, information about a user may be informed by the list of people that they are following. We performed feature extraction by encoding these networks as an adjacency matrix and apply singular-value decomposition (SVD) as a form of *dimensionality reduction* to reduce the feature dimension to a manageable size (specifically, 100) [12]. Like user features, social network features were included via simple concatenation.

## 3. Results and Discussion

Main results are presented in Table 1. Both linear and deep models performed well above random baselines (first two rows) in terms of *F*_β_. To facilitate comparison with a simple approach, in the third row we show the performance if we classify all tweeters whose tweets contain “Juul” as a substring as Juul users. Although precision is better than that for most other methods, recall is in single digits and lower compared to all other methods. Precision is more important, but this approach will drastically reduce the coverage we may be able to obtain. This is not surprising because rule-based methods well known to suffer from very low recall [13]. In contrast, in rows 6 and 7 of the table, we see precision close to the simple string matching-based method but much higher recall. Thus, we conclude that several Juul users may not be tweeting about Juul, but machine-learned models are a viable alternative for handling these cases.

**Table 1.** Our main results based on average performances from 5-fold cross-validation repeated six times. The ‘+’ symbol indicates the addition of a feature to the model with just the text features.

| **Model** | **$P$ (%)** | **$R$ (%)** | **$F_\beta$ (%)** | **$AP@10$ (%)** | **$P@10$ (%)** |
| --- | --- | --- | --- | --- | --- |
| Random Uniform | 25.79 | **51.10** | 26.57 | - | - |
| Random Stratified | 26.60 | 26.60 | 26.60 | - | - |
| Positive if “Juul” is in tweet text | 58.82 | 6.62 | 40.18 | - | - |
| Linear Model (Only text features) | 45.39 | 27.13 | 40.76 | 34.88 | 48.67 |
| + User features | 49.59 | 26.10 | 42.39 | 37.49 | 50.33 |
| + Friends network | **61.81** | 19.12 | **50.52** | 43.21 | 56.67 |
| + Followers network | 56.80 | 24.47 | 50.33 | **46.25** | **60.33** |
| Linear Model (All features) | 52.48 | 32.65 | 49.50 | 38.94 | 55.33 |
| Deep Model (Only text features) | 41.99 | 22.01 | 39.14 | 32.47 | 44.00 |
| Deep Model (All features) | 41.60 | 27.53 | 40.10 | 37.82 | 48.67 |

Between linear and deep models, the best linear model (with only text and friends/followers network features) performed at almost ten *F*_β_ and seven *AP*@10 points above the best deep neural model. Deep learning models (last two rows) seemed to overfit when trained on relatively small datasets and these results likely indicate that simpler models may be more suitable given the limited amount of training data. Features derived from the friends and followers network had the most substantial impact on performance such that we observe almost a 10-point improvement in *F*_β_ from simply adding either types of network feature. Follower features were slightly more beneficial than friend features; inclusion of these isolated features resulted in improvements from 34.88% (text-only features) to 46.25% (text and follower features) and 43.21% (text and friend features) on *AP*@10. Including more hand-crafted features generally improved recall without adversely affecting precision, and this is consistent for both linear and deep learning models. When evaluating holistically for a balance of both *F*_β_ and *AP*@10, the linear model with only follower-based features performed best at 50.33% *F*_β_ and 46.25% *AP*@10. A linear model that has all features proposed in the methods section lead to a recall of 32.65% with a precision of 52.48%, indicating the features are acting complementarily to improve recall with a comparable loss in precision. Given our precision focus, we believe this full model may not be worth exploring further. However, this model may be of utility for heavily funded intervention scenarios where the primary goal is to achieve better coverage at the expense of many false positives.

For interpretability, we additionally included the *P*@10 measure, which represents the precision rate evaluated on the top 10 instances when ranked by likelihood of a tweeter being a Juul user. Based on the main results, we achieved approximately 60% precision for the top 10 out of 118 examples (average number of examples per test fold). To illustrate a practical use case for the proposed model, we offer the following hypothetical scenario. Suppose there are 2000 individuals in the demographic for an intervention campaign — namely, first year students of a typical American university that regularly use social media. By simple extrapolation, approximately 169 students 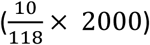 will be targeted in the campaign. With 60% precision, 101 of those targeted will be actual Juul users (169 × 0.6). As Juul use occurs with a prevalence of 25% according to our training data, a crude estimate of 500 students using Juul per 2000 would conclude that our model has recall of about 20% (101/500), which is consistent with our main results.

## 4 Conclusion and Future Work

In this study, we compared linear and deep learning models for automatically predicting whether an individual is positive or negative for Juul use (past 30 day use) based on their Twitter data, including tweets and social network features. Results indicate that linear models outperform deep learning models, and that inclusion of non-textual features, such as the network of friends and followers, is important for maximizing model performance. It is important to note that, given limited training data, it is not conclusive that linear models will always outperform deep learning models for this task. Future work will focus on curating a larger dataset; with an abundance of training data, we expect a substantial leap in performance for both linear and (especially) deep models. Moreover, future work will expand on these results with a more rigorous assessment and analysis of social network-based features by additionally incorporating the vast twitter feeds of an individual’s friends and followers (and not just the network itself) – we believe this will be especially helpful for predicting Juul use even among the many users with little or no social media activity. Other avenues of research include predictive modeling of more detailed usage patterns, nicotine dependency, and risk awareness. In terms of surveillance implications, our models can help public health campaigns selectively communicate preventative and tobacco treatment messages or clinical trial recruitment calls to potential users. Given not all Juul users are on Twitter (or other social media), we emphasize this is only one component of a potentially multi-pronged approach to reach the user space. However, this may be relatively inexpensive and can capture a segment of users that tend to actively use social media. As such, we believe this line of work may benefit future exploration by other members of the scientific community who work at the intersection of public health, informatics, and computer science.

## Data Availability

Not applicable

